## Supplementary for "Regional excess mortality during the 2020 COVID-19 pandemic: a study of five European countries"

### List of Figures

S1 NUTS2 (black) and NUTS3 (grey) region borders in Enlgand, Greece, Italy, Spain and Switzerland. 12

S2 Weekly trends for all-cause total number of deaths by gender and age in England. The blue curve represents the posterior mean predicted from the 2015–2019 model, while the shaded blue ribbon describes the posterior 95% interval. The black line shows the observed number of deaths for 2020. 13

S3 Weekly trends for all-cause total number of deaths by gender and age in Greece. The blue curve represents the posterior mean predicted from the 2015–2019 model, while the shaded blue ribbon describes the posterior 95% interval. The black line shows the observed number of deaths for 2020. 14

S4 Weekly trends for all-cause total number of deaths by gender and age in Italy. The blue curve represents the posterior mean predicted from the 2015–2019 model, while the shaded blue ribbon describes the posterior 95% interval. The black line shows the observed number of deaths for 2020. 15

S5 Weekly trends for all-cause total number of deaths by gender and age in Switzerland. The blue curve represents the posterior mean predicted from the 2015–2019 model, while the shaded blue ribbon describes the posterior 95% interval. The black line shows the observed number of deaths for 2020. . . . . . . . . . . . . . . . . . . . . . . . . . . . . . . . . . . . . . . . . . . . . 16

S6 Weekly trends for all-cause total number of deaths by gender and age in Switzerland. The blue curve represents the posterior mean predicted from the 2015–2019 model, while the shaded blue ribbon describes the posterior 95% interval. The black line shows the observed number of deaths for 2020. . . . . 17
S7 Relative excess mortality during 2020 by NUTS3 regions in England stratified by age and sex. . . 18
S8 Posterior probability that the relative excess mortality is larger than zero during 2020 by NUTS3 regions in England stratified by age and sex. . . . . 19
S9 Relative excess mortality during 2020 by NUTS3 regions in Greece stratified by age and sex. . . 20
S10 Posterior probability that the relative excess mortality is larger than zero during 2020 by NUTS3 regions in Greece stratified by age and sex. . . . . 21
S11 Relative excess mortality during 2020 by NUTS3 regions in Italy stratified by age and sex. . . . . 22
S12 Posterior probability that the relative excess mortality is larger than zero during 2020 by NUTS3 regions in Italy stratified by age and sex. . . . . 23
S13 Relative excess mortality during 2020 by NUTS3 regions in Spain stratified by age and sex. . . . 24
S14 Posterior probability that the relative excess mortality is larger than zero during 2020 by NUTS3 regions in Spain stratified by age and sex. . . . . 25
S15 Relative excess mortality during 2020 by NUTS3 regions in Switzerland stratified by age and sex. 26
S16 Posterior probability that the relative excess mortality is larger than zero during 2020 by NUTS3 regions in Switzerland stratified by age and sex. . . . . 27

Table S1: Cross-validation results: correlation between predicted and observed number of deaths, and 95% coverage probability across the different countries and age and sex groups.

| | England | | Greece | | Italy | | Spain | | Switzerland | |
|---|---|---|---|---|---|---|---|---|---|---|
| Age group | Correlation | Coverage | Correlation | Coverage | Correlation | Coverage | Correlation | Coverage | Correlation | Coverage |
| Males | | | | | | | | | | |
| $< 40$ | 0.23 (0.21, 0.24) | 0.94 | 0.56 (0.57, 0.62) | 0.92 | 0.51 (0.50, 0.52) | 0.94 | 0.69 (0.68, 0.70) | 0.93 | 0.41 (0.38, 0.44) | 0.93 |
| 40-59 | 0.45 (0.44, 0.46) | 0.95 | 0.87 (0.86, 0.87) | 0.94 | 0.83 (0.83, 0.84) | 0.95 | 0.90 (0.90, 0.91) | 0.94 | 0.69 (0.67, 0.71) | 0.94 |
| 60-69 | 0.55 (0.54, 0.55) | 0.95 | 0.90 (0.89, 0.90) | 0.95 | 0.87 (0.85, 0.88) | 0.95 | 0.92 (0.92, 0.93) | 0.94 | 0.74 (0.72, 0.76) | 0.94 |
| 70-79 | 0.70 (0.69, 0.71) | 0.95 | 0.93 (0.92, 0.93) | 0.95 | 0.93 (0.91, 0.93) | 0.95 | 0.95 (0.94, 0.95) | 0.93 | 0.84 (0.83, 0.85) | 0.95 |
| $geq$80 | 0.83 (0.82, 0.84) | 0.94 | 0.96 (0.95, 0.96) | 0.95 | 0.95 (0.94, 0.95) | 0.93 | 0.97 (0.97, 0.97) | 0.91 | 0.92 (0.91, 0.93) | 0.95 |
| Females | | | | | | | | | | |
| $40 <$ | 0.15 (0.14, 0.16) | 0.93 | 0.43 (0.40, 0.46) | 0.88 | 0.38 (0.37, 0.40) | 0.92 | 0.58 (0.56, 0.59) | 0.92 | 0.28 (0.26, 0.31) | 0.89 |
| 40-59 | 0.35 (0.34, 0.36) | 0.95 | 0.81 (0.79, 0.81) | 0.93 | 0.78 (0.77, 0.79) | 0.95 | 0.86 (0.85, 0.86) | 0.94 | 0.57 (0.55, 0.59) | 0.93 |
| 60-69 | 0.46 (0.45, 0.47) | 0.95 | 0.86 (0.85, 0.86) | 0.94 | 0.83 (0.82, 0.83) | 0.95 | 0.88 (0.87, 0.88) | 0.95 | 0.64 (0.63, 0.66) | 0.94 |
| 70-79 | 0.65 (0.63, 0.65) | 0.95 | 0.92 (0.91, 0.92) | 0.95 | 0.91 (0.90, 0.92) | 0.95 | 0.92 (0.91, 0.93) | 0.94 | 0.81 (0.80, 0.82) | 0.94 |
| $geq$80 | 0.87 (0.82, 0.84) | 0.94 | 0.97 (0.96, 0.97) | 0.94 | 0.96 (0.95, 0.96) | 0.92 | 0.97 (0.97, 0.98) | 0.90 | 0.94 (0.93, 0.95) | 0.94 |

Table S2: Median relative excess deaths and 95% credible intervals in 2020 per NUTS2 regions in England.

| Region | Males | Females |
|---|---|---|
| Outer London - West and North West | 0.21 (0.15, 0.28) | 0.12 (0.05, 0.28) |
| Outer London - East and North East | 0.20 (0.14, 0.26) | 0.15 (0.07, 0.26) |
| West Midlands | 0.19 (0.13, 0.25) | 0.14 (0.06, 0.25) |
| Inner London - East | 0.19 (0.13, 0.26) | 0.15 (0.08, 0.26) |
| Outer London - South | 0.16 (0.10, 0.24) | 0.08 (-0.00, 0.24) |
| South Yorkshire | 0.16 (0.10, 0.22) | 0.12 (0.05, 0.22) |
| Lancashire | 0.15 (0.09, 0.21) | 0.08 (0.01, 0.21) |
| Greater Manchester | 0.14 (0.09, 0.20) | 0.09 (0.02, 0.20) |
| West Yorkshire | 0.14 (0.08, 0.20) | 0.08 (0.01, 0.20) |
| Kent | 0.14 (0.07, 0.20) | 0.08 (0.01, 0.20) |
| Merseyside | 0.13 (0.07, 0.19) | 0.10 (0.02, 0.19) |
| Leicestershire, Rutland and Northamptonshire | 0.13 (0.07, 0.19) | 0.07 (-0.01, 0.19) |
| Bedfordshire and Hertfordshire | 0.12 (0.06, 0.18) | 0.08 (0.00, 0.18) |
| Shropshire and Staffordshire | 0.11 (0.06, 0.18) | 0.07 (0.00, 0.18) |
| Tees Valley and Durham | 0.11 (0.05, 0.17) | 0.09 (0.00, 0.17) |
| Essex | 0.11 (0.05, 0.17) | 0.03 (-0.04, 0.17) |
| Cumbria | 0.10 (0.03, 0.17) | 0.06 (-0.02, 0.17) |
| Herefordshire, Worcestershire and Warwickshire | 0.10 (0.04, 0.16) | 0.07 (-0.00, 0.16) |
| East Yorkshire and Northern Lincolnshire | 0.09 (0.03, 0.16) | 0.04 (-0.03, 0.16) |
| Inner London - West | 0.09 (0.03, 0.16) | 0.02 (-0.06, 0.16) |
| Derbyshire and Nottinghamshire | 0.09 (0.04, 0.15) | 0.04 (-0.04, 0.15) |
| Lincolnshire | 0.09 (0.03, 0.16) | 0.06 (-0.02, 0.16) |
| Northumberland and Tyne and Wear | 0.08 (0.03, 0.14) | 0.05 (-0.02, 0.14) |
| Berkshire, Buckinghamshire and Oxfordshire | 0.08 (0.02, 0.14) | 0.06 (-0.02, 0.14) |
| Surrey, East and West Sussex | 0.07 (0.01, 0.12) | 0.05 (-0.03, 0.12) |
| Cheshire | 0.07 (0.01, 0.13) | 0.06 (-0.02, 0.13) |
| Gloucestershire, Wiltshire and Bath/Bristol area | 0.06 (0.01, 0.12) | 0.04 (-0.03, 0.12) |
| Hampshire and Isle of Wight | 0.04 (-0.01, 0.10) | 0.02 (-0.05, 0.10) |
| East Anglia | 0.04 (-0.01, 0.09) | 0.03 (-0.05, 0.09) |
| North Yorkshire | 0.04 (-0.02, 0.10) | 0.03 (-0.05, 0.10) |
| Dorset and Somerset | 0.02 (-0.03, 0.08) | -0.00 (-0.08, 0.08) |
| Cornwall and Isles of Scilly | -0.02 (-0.08, 0.04) | -0.05 (-0.11, 0.04) |
| Devon | -0.03 (-0.08, 0.03) | -0.05 (-0.12, 0.03) |
| England | 0.11 (0.05, 0.16) | 0.06 (-0.01, 0.16) |

Table S3: Median relative excess deaths and 95% credible intervals in 2020 per NUTS2 regions in Greece.

| Region | Males | Females |
|---|---|---|
| Eastern Macedonia and Thrace | 0.17 (0.09, 0.25) | 0.14 (0.06, 0.25) |
| Western Macedonia | 0.16 (0.08, 0.27) | 0.19 (0.08, 0.27) |
| Central Macedonia | 0.15 (0.08, 0.24) | 0.13 (0.05, 0.24) |
| Thessaly | 0.08 (0.01, 0.17) | 0.09 (0.01, 0.17) |
| Western Greece | 0.05 (-0.02, 0.12) | 0.01 (-0.07, 0.12) |
| Ionian Islands | 0.04 (-0.04, 0.13) | 0.01 (-0.08, 0.13) |
| Attica | 0.02 (-0.04, 0.09) | 0.02 (-0.05, 0.09) |
| Central Greece | 0.02 (-0.05, 0.10) | -0.00 (-0.08, 0.10) |
| Epirus | -0.00 (-0.08, 0.09) | 0.01 (-0.07, 0.09) |
| Peloponnese | -0.00 (-0.07, 0.08) | 0.05 (-0.04, 0.08) |
| Crete | -0.01 (-0.08, 0.06) | 0.02 (-0.06, 0.06) |
| North Aegean | -0.01 (-0.10, 0.08) | 0.06 (-0.04, 0.08) |
| South Aegean | -0.03 (-0.10, 0.05) | 0.02 (-0.06, 0.05) |
| Greece | 0.06 (-0.01, 0.13) | 0.06 (-0.02, 0.13) |

Table S4: Median relative excess deaths and 95% credible intervals in 2020 per NUTS2 regions in Italy.

| Region | Males | Females |
|---|---|---|
| Lombardia | 0.29 (0.20, 0.38) | 0.25 (0.17, 0.38) |
| Piemonte | 0.18 (0.10, 0.27) | 0.17 (0.08, 0.27) |
| Trentino-Alto Adige | 0.16 (0.08, 0.25) | 0.20 (0.11, 0.25) |
| Valle d'Aosta | 0.15 (0.05, 0.28) | 0.23 (0.10, 0.28) |
| Liguria | 0.15 (0.06, 0.23) | 0.12 (0.04, 0.23) |
| Emilia-Romagna | 0.13 (0.04, 0.21) | 0.12 (0.04, 0.21) |
| Veneto | 0.10 (0.02, 0.17) | 0.11 (0.02, 0.17) |
| Marche | 0.07 (-0.01, 0.15) | 0.08 (0.01, 0.15) |
| Puglia | 0.07 (-0.00, 0.14) | 0.05 (-0.02, 0.14) |
| Friuli Venezia Giulia | 0.06 (-0.02, 0.13) | 0.08 (-0.00, 0.13) |
| Sardegna | 0.05 (-0.02, 0.12) | 0.06 (-0.02, 0.12) |
| Abruzzo | 0.04 (-0.04, 0.11) | 0.01 (-0.06, 0.11) |
| Toscana | 0.03 (-0.04, 0.11) | 0.04 (-0.03, 0.11) |
| Campania | 0.03 (-0.03, 0.10) | -0.00 (-0.07, 0.10) |
| Sicilia | 0.02 (-0.05, 0.09) | 0.02 (-0.05, 0.09) |
| Basilicata | 0.02 (-0.05, 0.10) | 0.03 (-0.06, 0.10) |
| Umbria | 0.01 (-0.06, 0.09) | 0.01 (-0.07, 0.09) |
| Calabria | 0.01 (-0.06, 0.08) | 0.00 (-0.07, 0.08) |
| Lazio | 0.00 (-0.07, 0.06) | -0.02 (-0.09, 0.06) |
| Molise | -0.00 (-0.08, 0.08) | 0.06 (-0.03, 0.08) |
| Italy | 0.10 (0.03, 0.18) | 0.09 (0.02, 0.18) |

Table S5: Median relative excess deaths and 95% credible intervals in 2020 per NUTS2 regions in Spain.

| Region | Males | Females |
|---|---|---|
| Madrid | 0.33 (0.27, 0.39) | 0.28 (0.20, 0.35) |
| Castile-la Mancha | 0.32 (0.26, 0.38) | 0.32 (0.23, 0.40) |
| Castile-leon | 0.28 (0.22, 0.34) | 0.28 (0.20, 0.36) |
| La Rioja | 0.18 (0.11, 0.28) | 0.16 (0.06, 0.26) |
| Catalonia | 0.15 (0.09, 0.20) | 0.18 (0.10, 0.25) |
| Aragon | 0.15 (0.09, 0.21) | 0.20 (0.12, 0.28) |
| Extremadura | 0.12 (0.06, 0.18) | 0.15 (0.07, 0.23) |
| Ceuta | 0.11 (-0.02, 0.30) | 0.31 (0.14, 0.54) |
| Navarre | 0.10 (0.04, 0.17) | 0.15 (0.07, 0.23) |
| Principality Of Asturias | 0.08 (0.03, 0.13) | 0.10 (0.03, 0.16) |
| Basque Community | 0.07 (0.02, 0.12) | 0.09 (0.02, 0.16) |
| Region Of Murcia | 0.04 (-0.01, 0.10) | 0.02 (-0.04, 0.09) |
| Cantabria | 0.04 (-0.02, 0.10) | 0.06 (-0.01, 0.14) |
| Andalusia | 0.03 (-0.01, 0.08) | 0.04 (-0.02, 0.10) |
| Valencian Community | 0.03 (-0.02, 0.07) | 0.02 (-0.04, 0.08) |
| Melilla | -0.01 (-0.13, 0.16) | 0.13 (-0.01, 0.30) |
| Galicia | -0.02 (-0.06, 0.03) | 0.01 (-0.06, 0.07) |
| Canary Islands | -0.08 (-0.12, -0.03) | -0.05 (-0.10, 0.01) |
| Balearic Islands | -0.08 (-0.13, -0.04) | -0.07 (-0.13, -0.01) |
| Spain | 0.12 (0.06, 0.19) | 0.12 (0.06, 0.19) |

Table S6: Median relative excess deaths and 95% credible intervals in 2020 per NUTS2 regions in Switzerland.

| Region | Males | Females |
|---|---|---|
| Ticino | 0.20 (0.11, 0.32) | 0.18 (0.09, 0.32) |
| Lake Geneva region | 0.16 (0.08, 0.25) | 0.14 (0.07, 0.25) |
| Eastern Switzerland | 0.08 (0.00, 0.16) | 0.07 (0.01, 0.16) |
| Espace Mittelland | 0.08 (0.00, 0.16) | 0.05 (-0.01, 0.16) |
| Northwestern Switzerland | 0.06 (-0.01, 0.15) | 0.04 (-0.02, 0.15) |
| Central Switzerland | 0.05 (-0.02, 0.13) | 0.01 (-0.05, 0.13) |
| Zurich | 0.02 (-0.06, 0.10) | 0.05 (-0.01, 0.10) |
| Switzerland | 0.08 (0.01, 0.16) | 0.07 (0.01, 0.16) |

Table S7: Frequency of population by age and sex groups in England, Greece, Italy, Spain and Switzerland in 2019.

| Age | Sex | England | Greece | Italy | Spain | Switzerland |
|---|---|---|---|---|---|---|
| 40< | female | 0.25 | 0.23 | 0.19 | 0.21 | 0.23 |
| 40< | male | 0.25 | 0.23 | 0.20 | 0.22 | 0.24 |
| 40-59 | female | 0.13 | 0.15 | 0.16 | 0.16 | 0.14 |
| 40-59 | male | 0.13 | 0.14 | 0.15 | 0.16 | 0.15 |
| 60-69 | female | . 0.05 | 0.06 | 0.06 | 0.06 | 0.06 |
| 60-69 | male | 0.05 | 0.06 | 0.06 | 0.05 | 0.05 |
| 70-79 | female | 0.04 | 0.05 | 0.05 | 0.04 | 0.04 |
| 70-79 | male | 0.04 | 0.04 | 0.05 | 0.04 | 0.04 |
| 80+ | female | . 0.03 | 0.02 | 0.05 | 0.04 | 0.03 |
| 80+ | male | 0.02 | 0.02 | 0.03 | 0.02 | 0.02 |

Table S8: Data sources for deaths and population across the different countries.

| country | Deaths | Population | Time point |
|---|---|---|---|
| England | Small Area Health Statistics Unit (SAHSU) | Office for National Statistics (ONS) | 30-06 |
| Greece | Hellenic Statistical Authority (ELSTAT) | ELSTAT | 01-01 |
| Italy | Italian National Institute of Statistics (ISTAT) | ISTAT | 01-01 |
| Spain | National Statistics Institute (INE) | INE | 01-01 |
| Switzerland | Federal Statistical Office (BFS) | BFS | 01-01 |

Fig. S1: NUTS2 (black) and NUTS3 (grey) region borders in Enlgand, Greece, Italy, Spain and Switzerland.

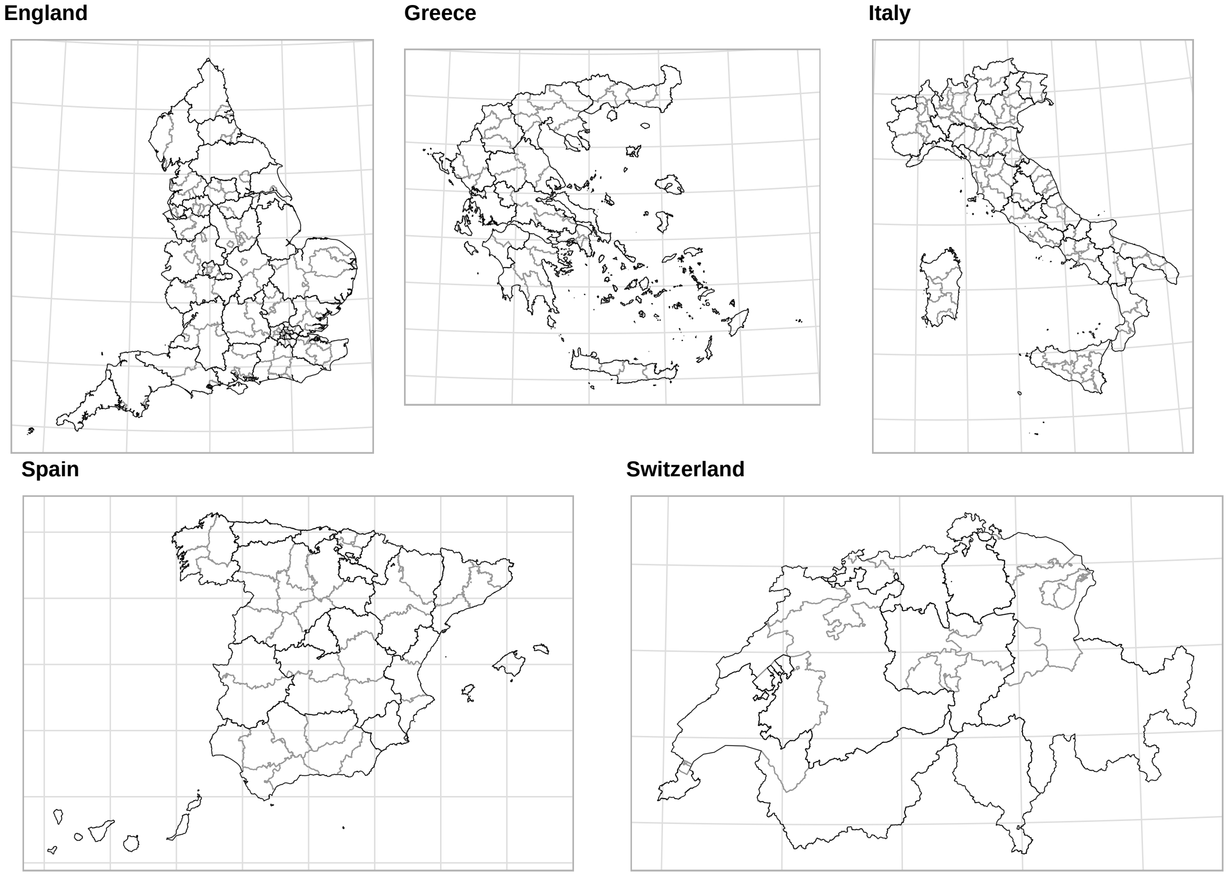

Fig. S2: Weekly trends for all-cause total number of deaths by gender and age in England. The blue curve represents the posterior mean predicted from the 2015–2019 model, while the shaded blue ribbon describes the posterior 95% interval. The black line shows the observed number of deaths for 2020.

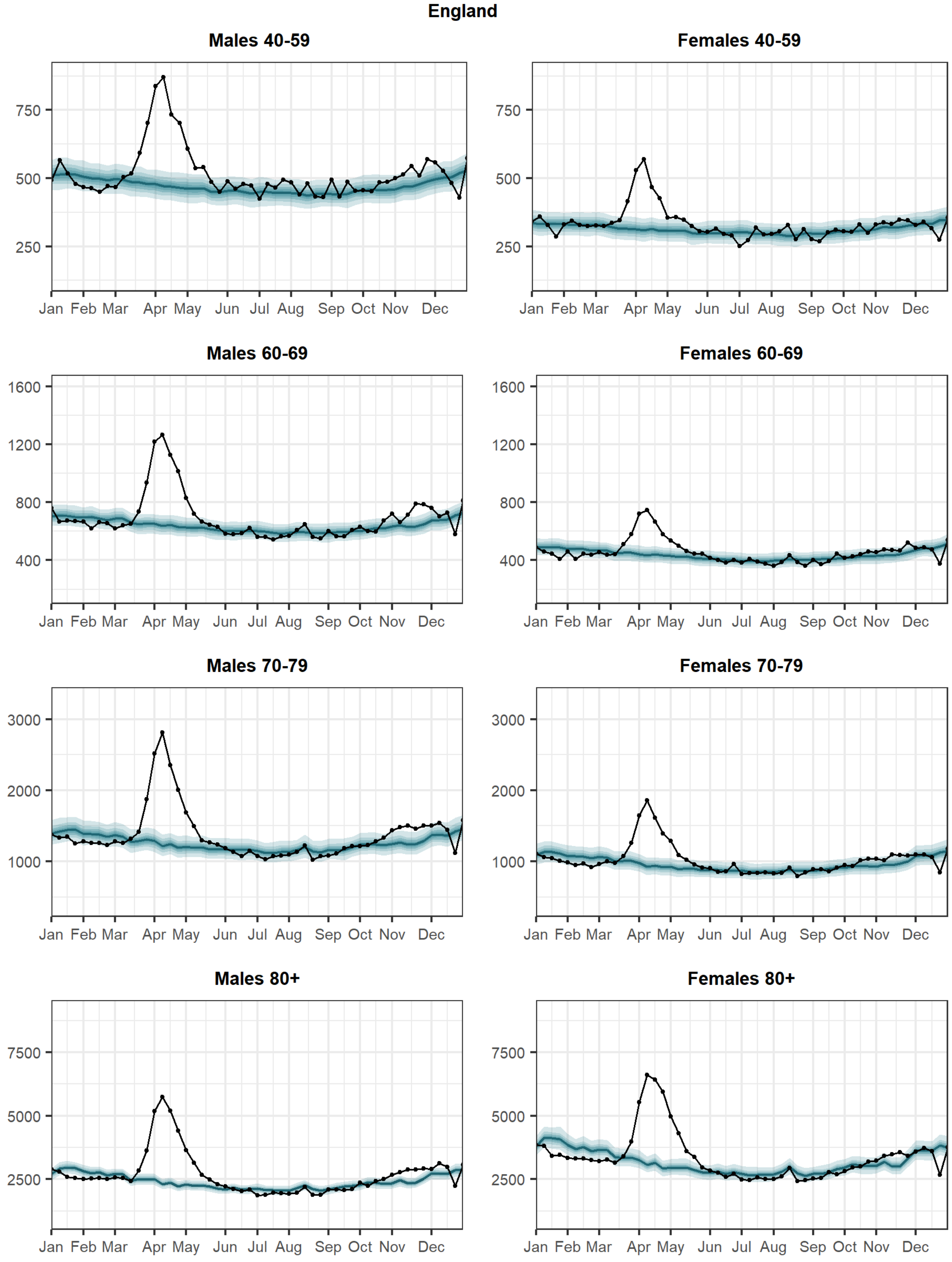

Fig. S3: Weekly trends for all-cause total number of deaths by gender and age in Greece. The blue curve represents the posterior mean predicted from the 2015–2019 model, while the shaded blue ribbon describes the posterior 95% interval. The black line shows the observed number of deaths for 2020.

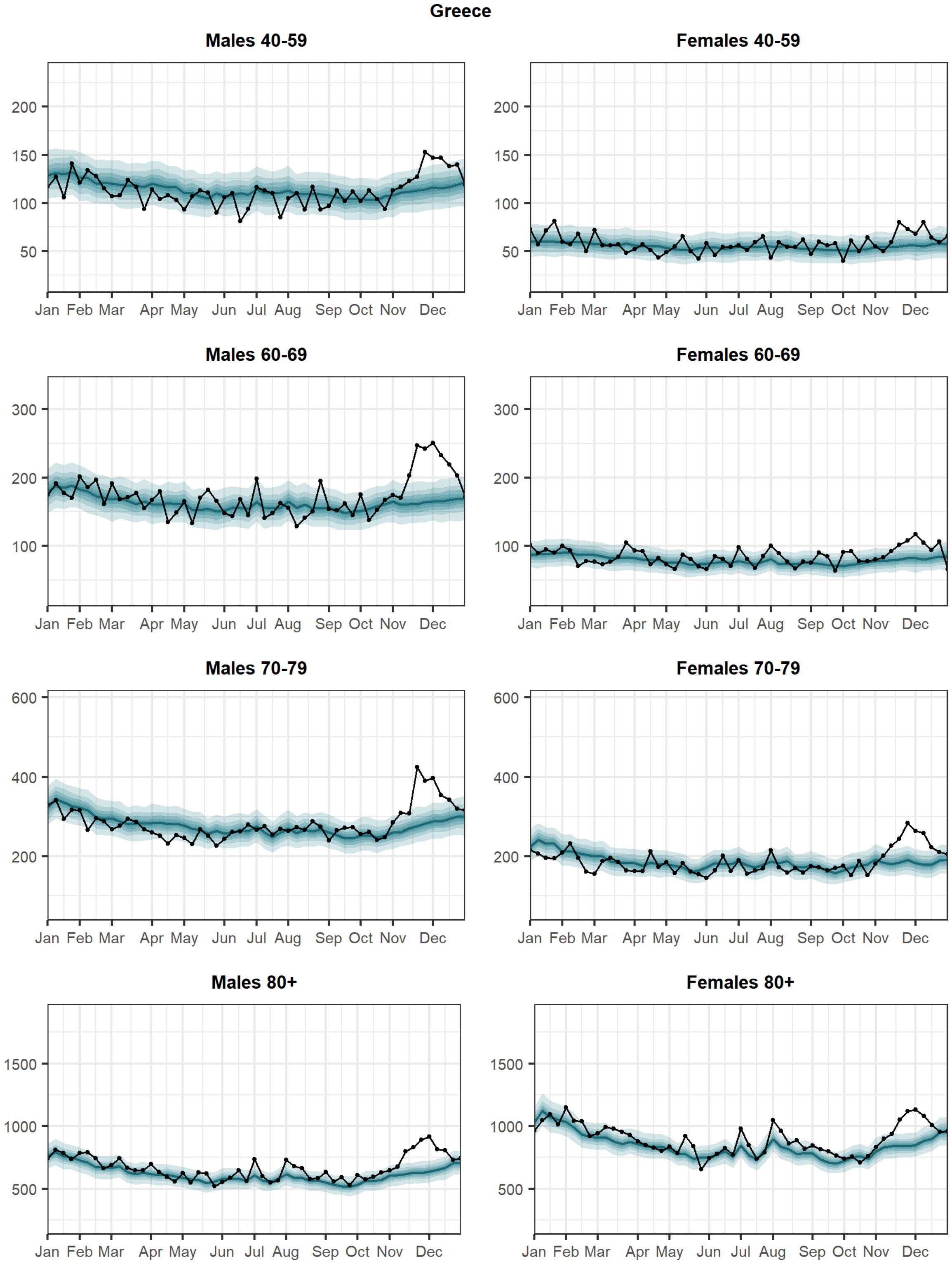

Fig. S4: Weekly trends for all-cause total number of deaths by gender and age in Italy. The blue curve represents the posterior mean predicted from the 2015–2019 model, while the shaded blue ribbon describes the posterior 95% interval. The black line shows the observed number of deaths for 2020.

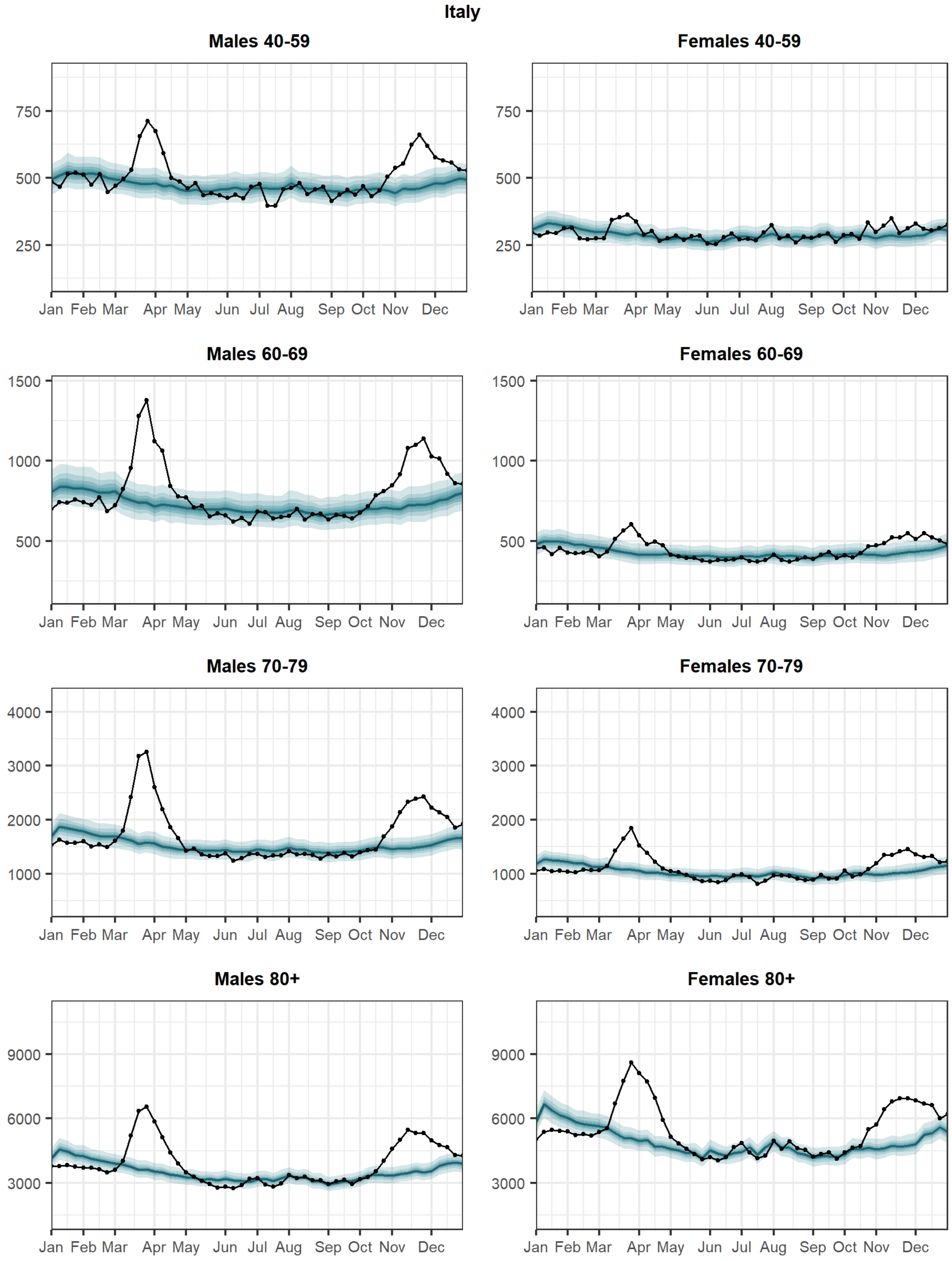

Fig. S5: Weekly trends for all-cause total number of deaths by gender and age in Switzerland. The blue curve represents the posterior mean predicted from the 2015–2019 model, while the shaded blue ribbon describes the posterior 95% interval. The black line shows the observed number of deaths for 2020.

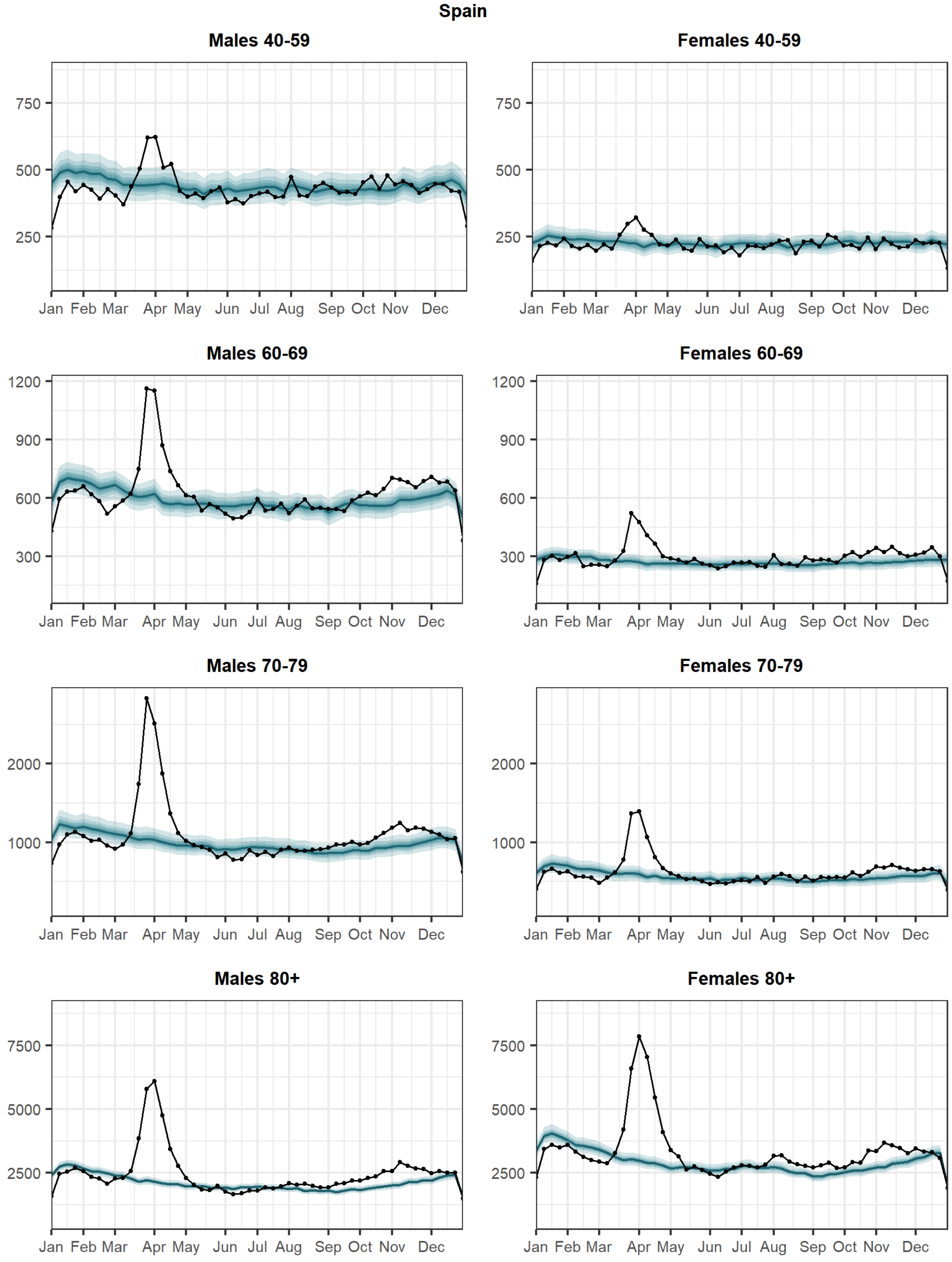

Fig. S6: Weekly trends for all-cause total number of deaths by gender and age in Switzerland. The blue curve represents the posterior mean predicted from the 2015–2019 model, while the shaded blue ribbon describes the posterior 95% interval. The black line shows the observed number of deaths for 2020.

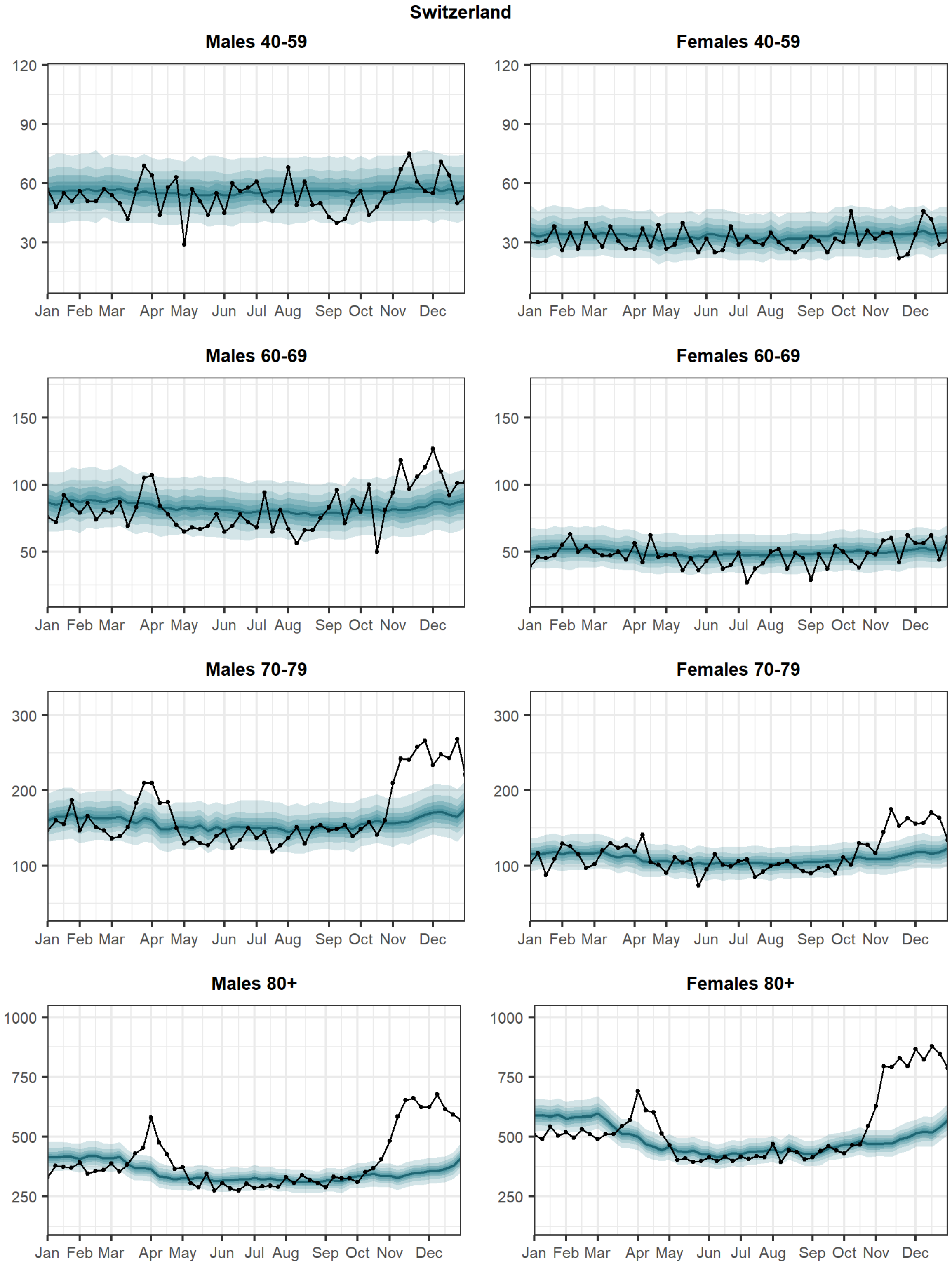

Fig. S7: Relative excess mortality during 2020 by NUTS3 regions in England stratified by age and sex.

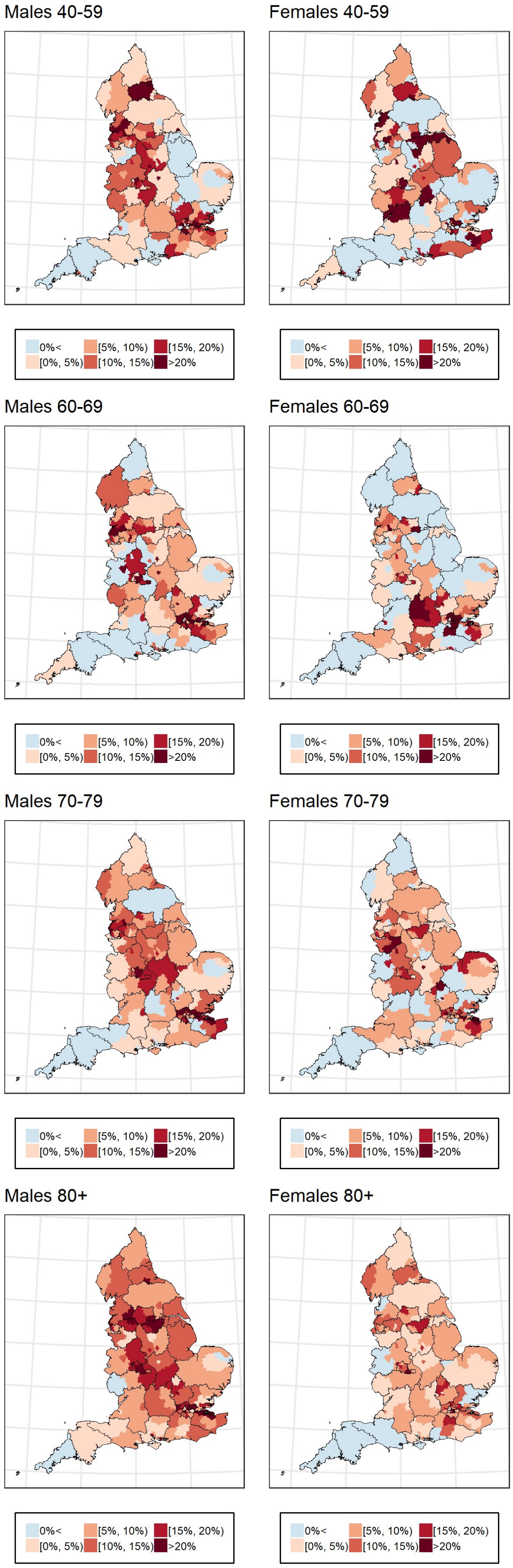

Fig. S8: Posterior probability that the relative excess mortality is larger than zero during 2020 by NUTS3 regions in England stratified by age and sex.

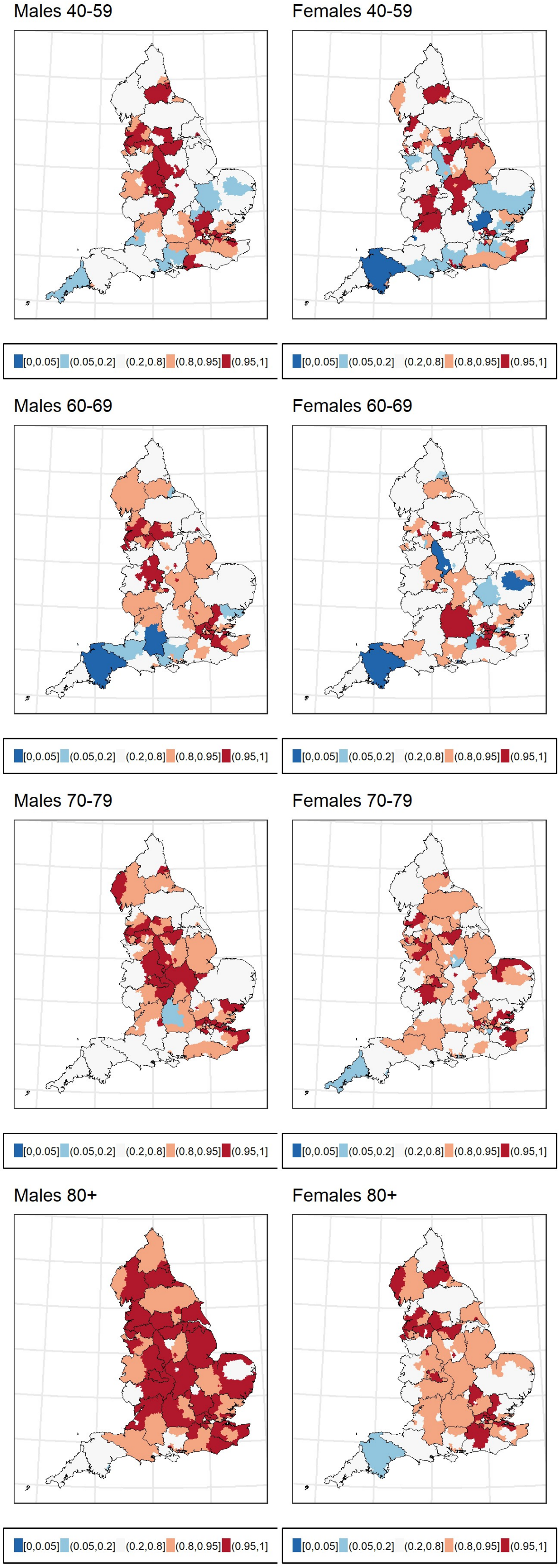

Fig. S9: Relative excess mortality during 2020 by NUTS3 regions in Greece stratified by age and sex.

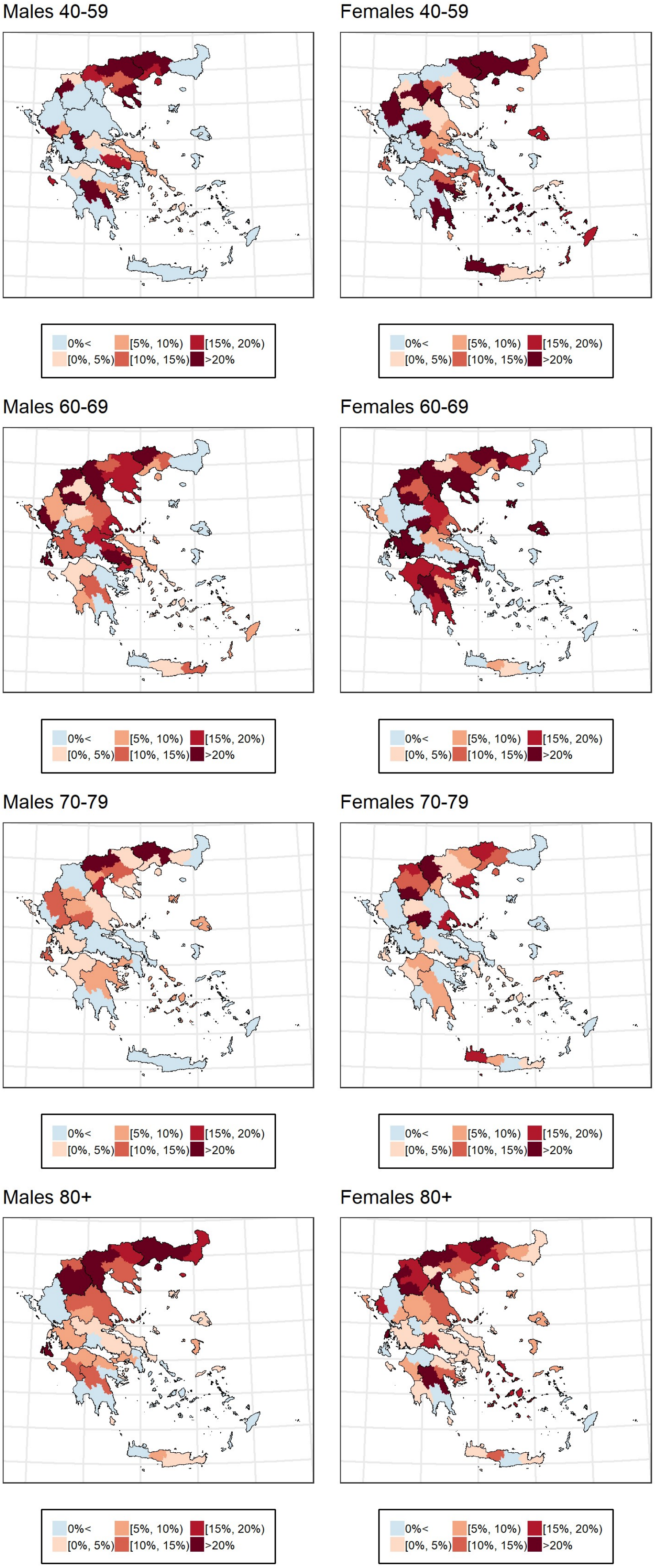

Fig. S10: Posterior probability that the relative excess mortality is larger than zero during 2020 by NUTS3 regions in Greece stratified by age and sex.

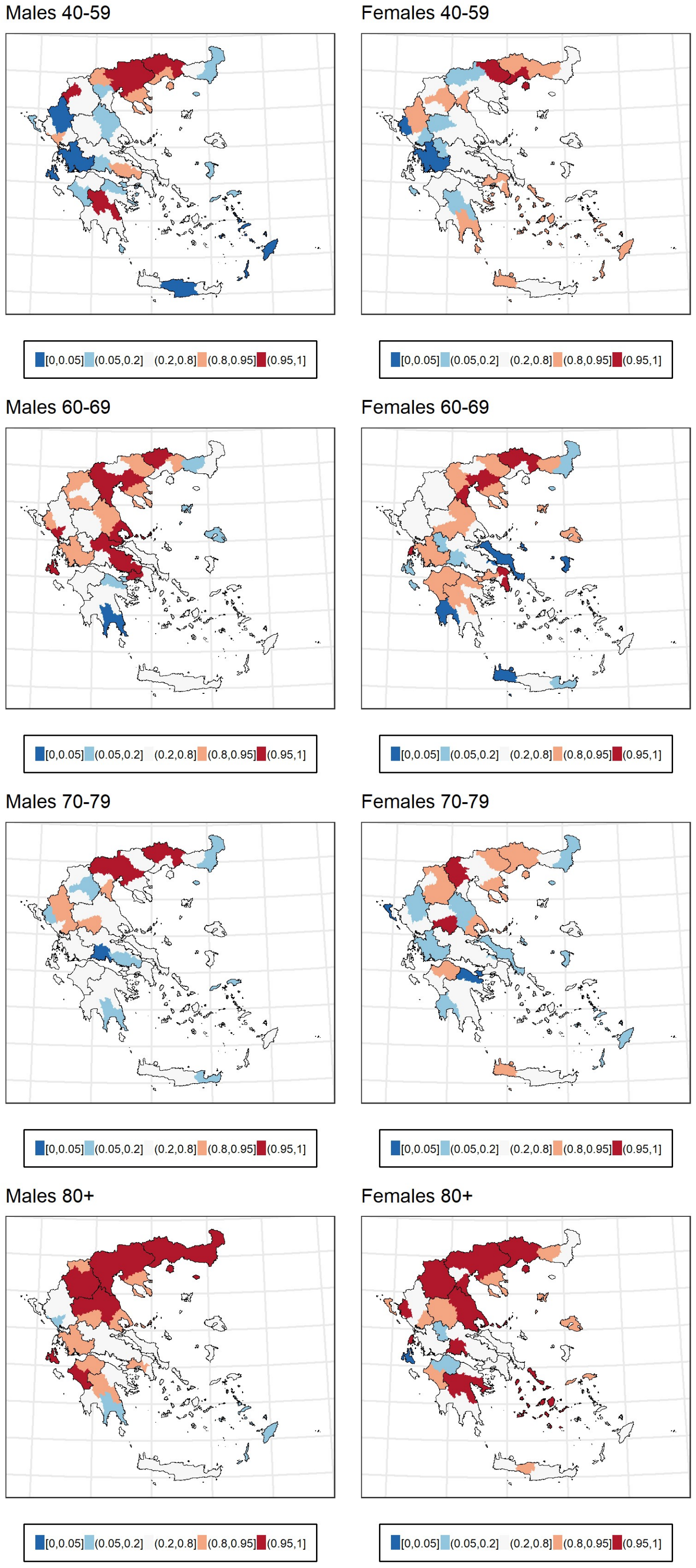

Fig. S11: Relative excess mortality during 2020 by NUTS3 regions in Italy stratified by age and sex.

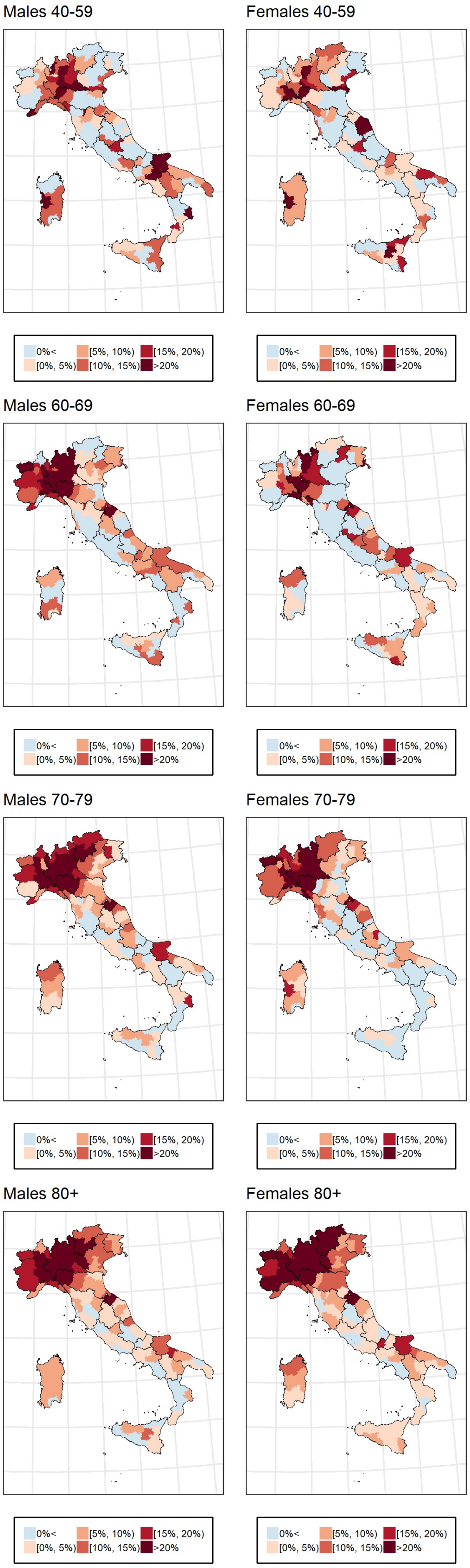

Fig. S12: Posterior probability that the relative excess mortality is larger than zero during 2020 by NUTS3 regions in Italy stratified by age and sex.

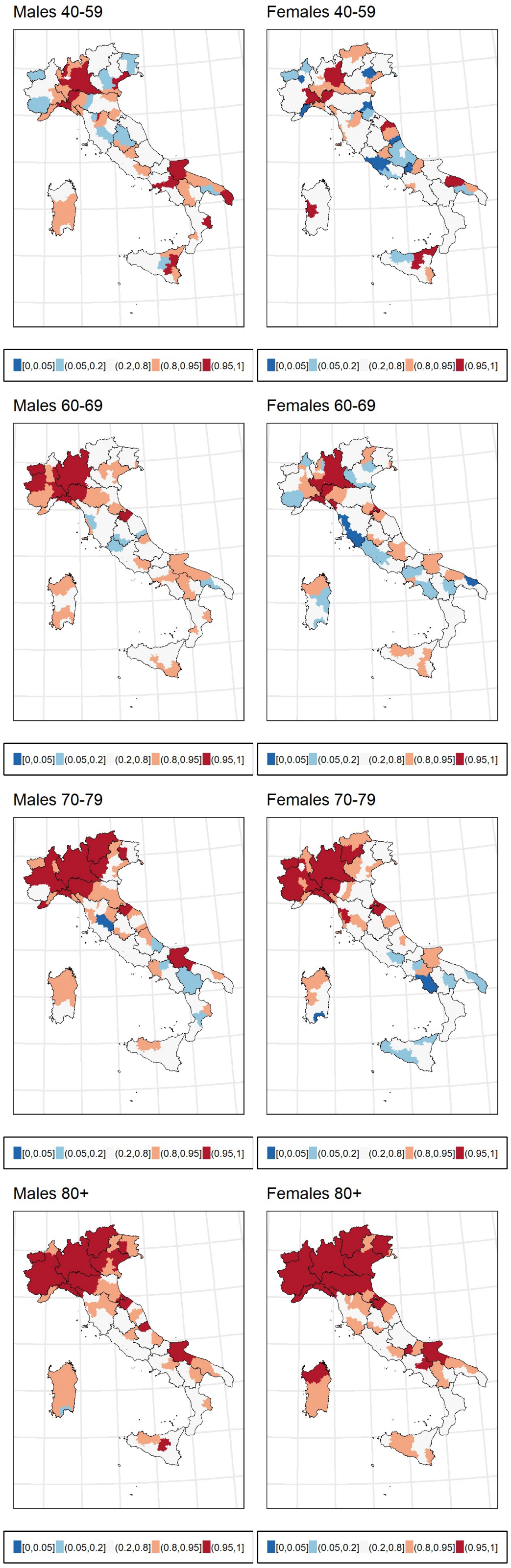

Fig. S13: Relative excess mortality during 2020 by NUTS3 regions in Spain stratified by age and sex.

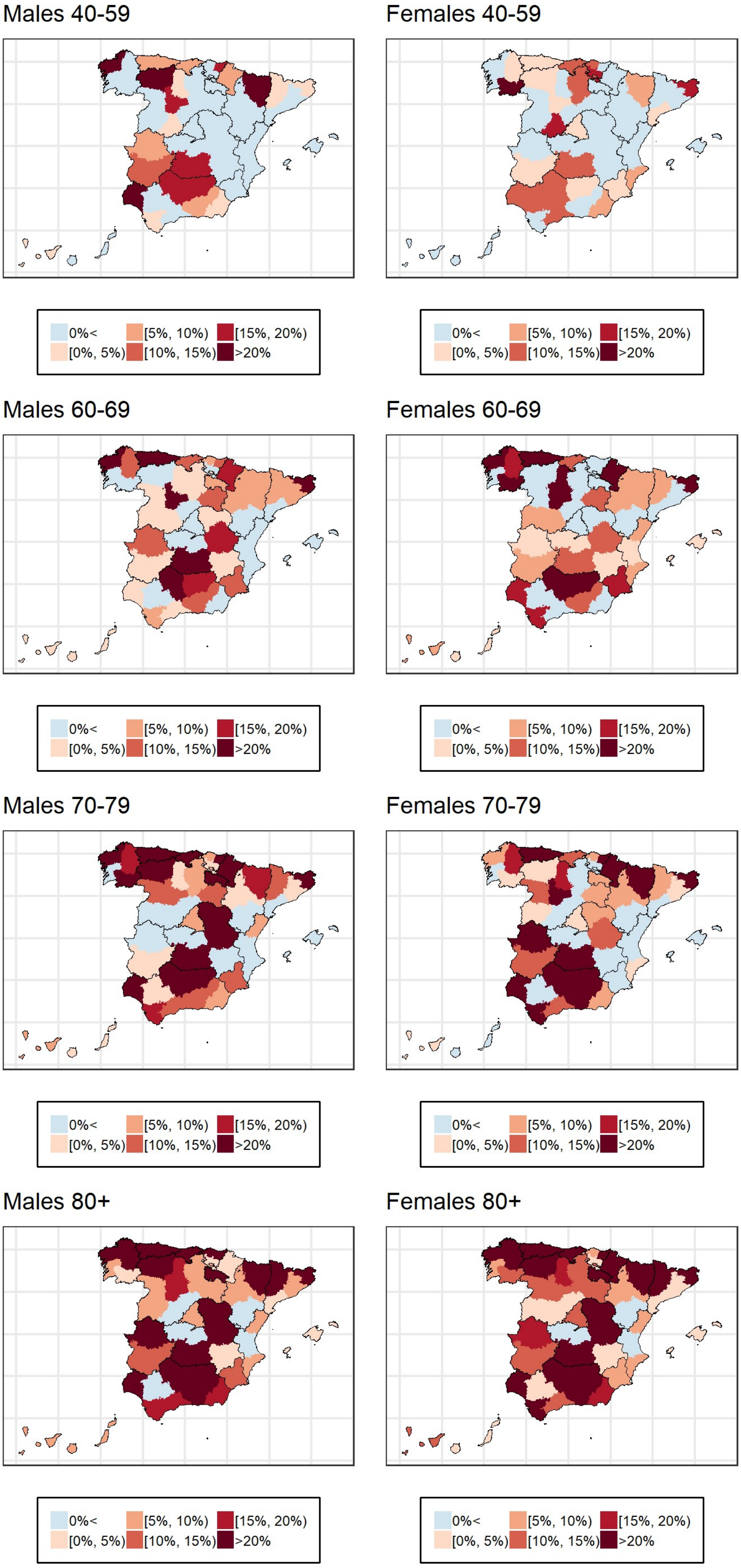

Fig. S14: Posterior probability that the relative excess mortality is larger than zero during 2020 by NUTS3 regions in Spain stratified by age and sex.

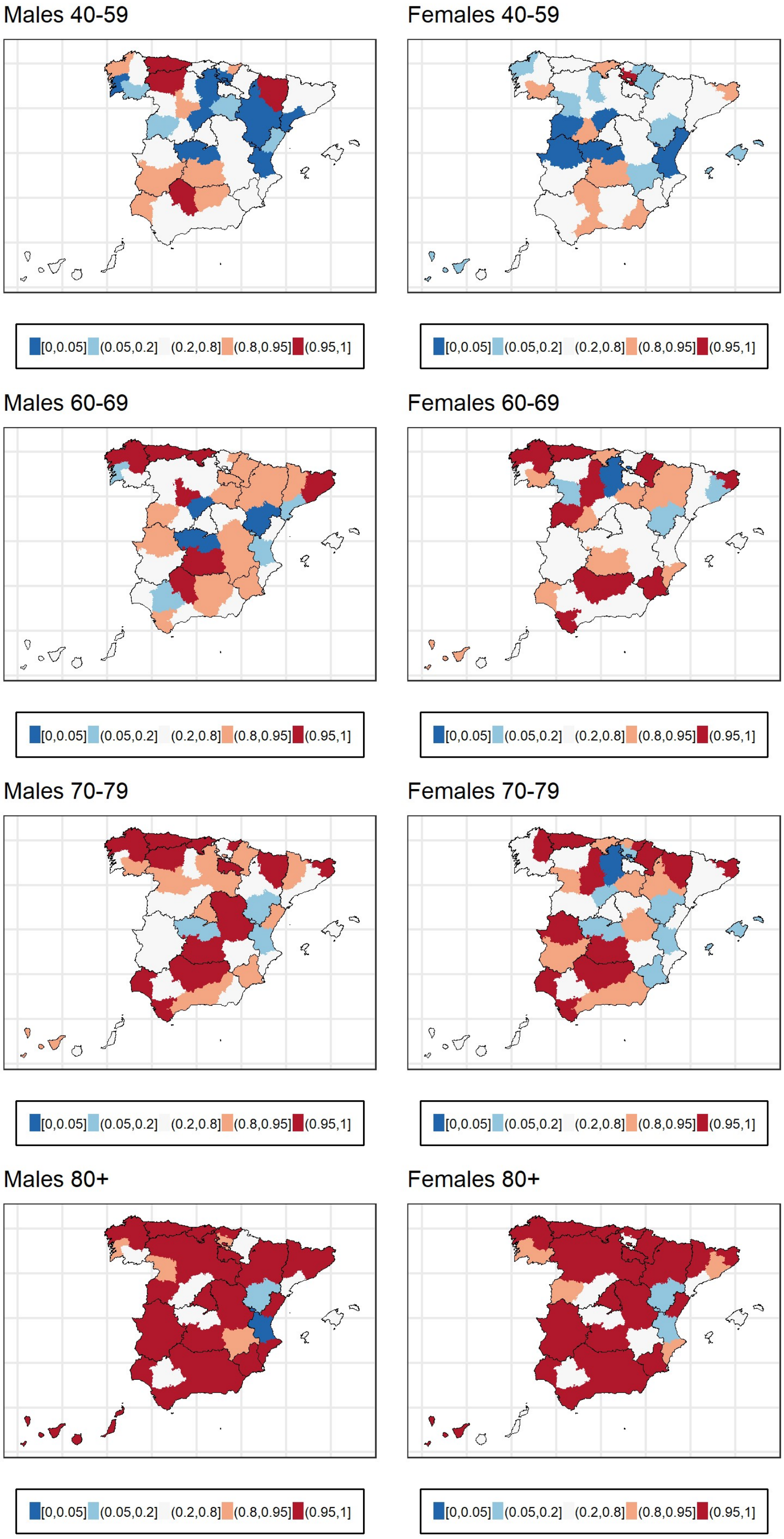

Fig. S15: Relative excess mortality during 2020 by NUTS3 regions in Switzerland stratified by age and sex.

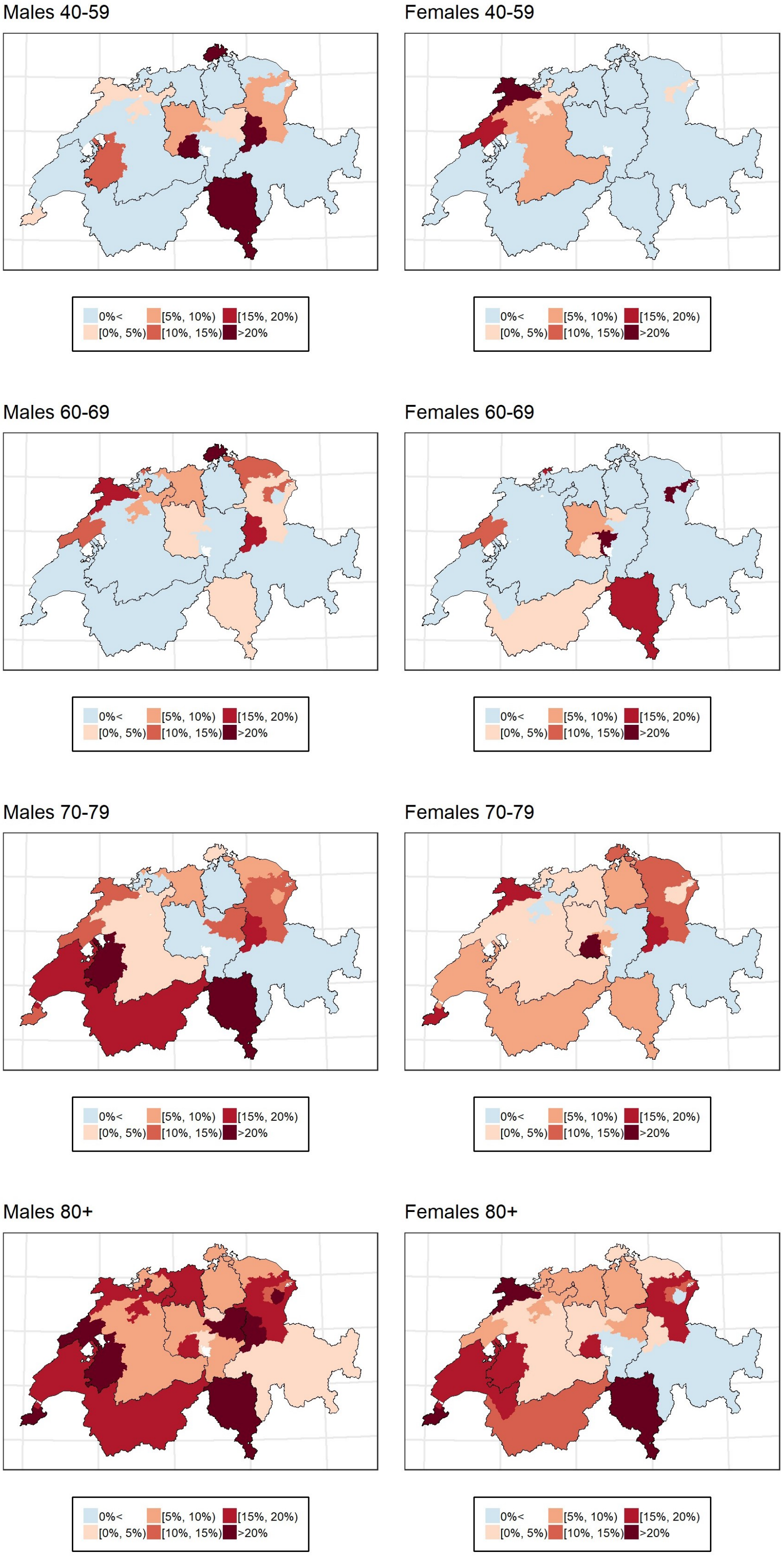

Fig. S16: Posterior probability that the relative excess mortality is larger than zero during 2020 by NUTS3 regions in Switzerland stratified by age and sex.

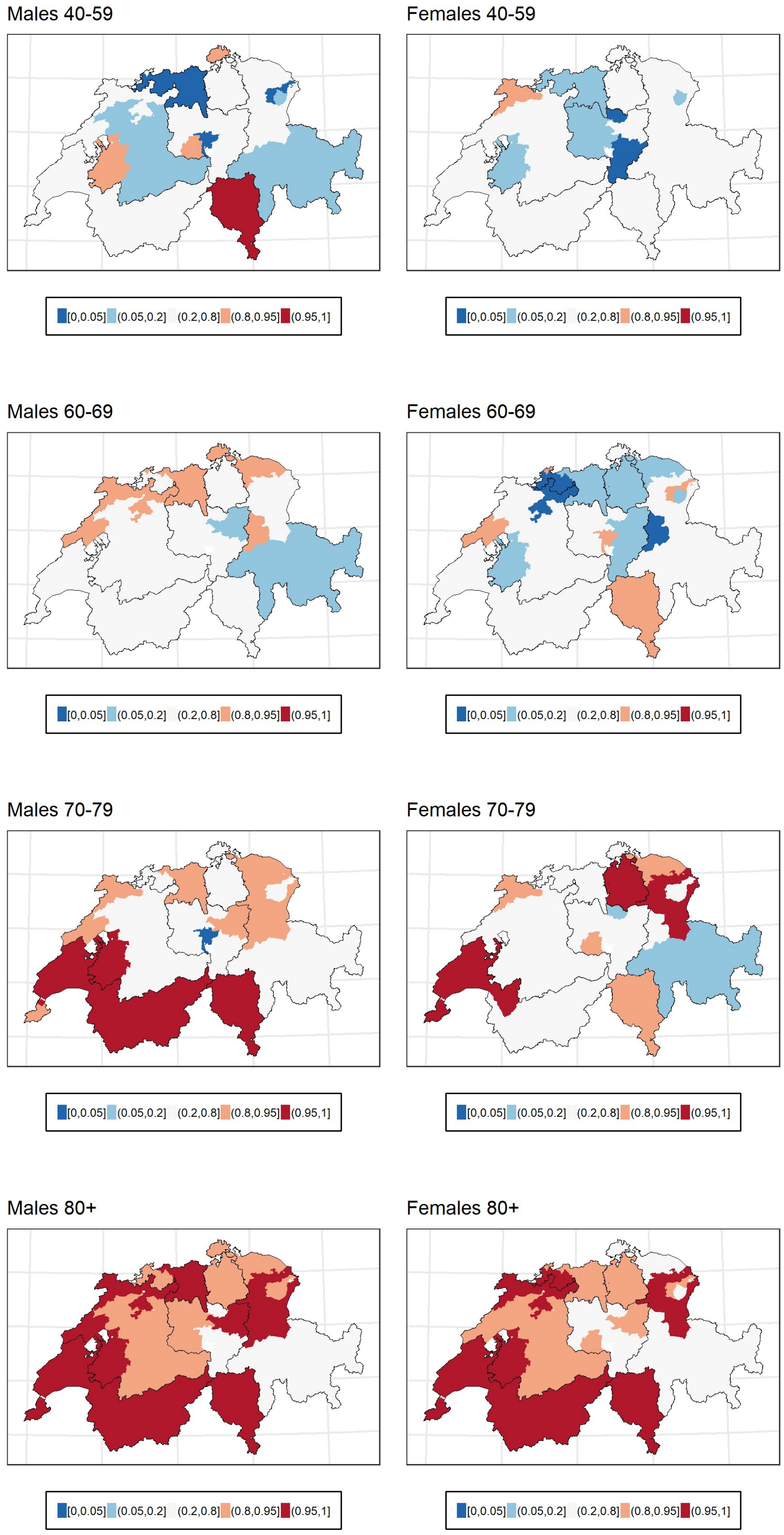
